# The impact of early life experiences and gut microbiota on neurobehavioral development among preterm infants: A longitudinal cohort study

**DOI:** 10.1101/2023.01.04.23284200

**Authors:** Jie Chen, Hongfei Li, Tingting Zhao, Kun Chen, Ming-Hui Chen, Zhe Sun, Wanli Xu, Kendra Maas, Barry Lester, Xiaomei Cong

## Abstract

**Objectives:** To investigate the impact of early life experiences and gut microbiota on neurobehavioral development among preterm infants during neonatal intensive care unit (NICU) hospitalization.

**Methods:** Preterm infants were followed from the NICU admission until their 28^th^ postnatal day or until discharge. Daily stool samples, painful/stressful experiences, feeding patterns, and other clinical and demographic data were collected. Gut microbiota was profiled using 16S rRNA sequencing, and operational taxonomic units (OTUs) were selected to predict the neurobehaviors. The neurobehavioral development was assessed by the Neonatal Neurobehavioral Scale (NNNS) at 36 to 38 weeks of post-menstrual age (PMA). Fifty-five infants who had NNNS measurements were included in the sparse log-contrast regression analysis.

**Results:** Preterm infants who experienced high level of pain/stress during the NICU hospitalization that were associated with higher NNNS stress/abstinence scores. Eight operational taxonomic units (OTUs) were identified to be associated with of NNNS subscales after controlling demographic and clinical features, feeding patterns, and painful/stressful experiences. These OTUs, taxa belong to seven genera including *Enterobacteriaceae_unclassified, Escherichia-Shigella, Incertae_Sedis, Veillonella, Enterococcus, Clostridium_sensu_stricto_1*, and *Streptococcus* with five belonging to *Firmicutes* and two belonging to *Proteobacteria* phylum. The enriched abundance of *Enterobacteriaceae_unclassified* (OTU17) and *Streptococcus* (OTU28) were consistently associated with less optimal neurobehavioral outcomes. The other six OTUs were also associated with infant neurobehavioral responses depending on days at NICU stay.

**Conclusions:** This study explored the dynamic impact of specific OTUs on neurobehavioral development among preterm infants after controlling for early life experiences, i.e., acute and chronic pain/stress, and feeding in the NICU.

## 1. Introduction

The mortality rate of preterm infants has significantly decreased recent years along with the advances of neonatal healthcare and medical treatments (Swamy et al., 2008; Glass et al., 2015; Bell et al., 2022), whereas preterm infants are still at high risk of neurodevelopmental deficiency in early life as well as late childhood mortality, and late onset mental and behavioral disorders (Vohr, 2013; Patel, 2016; Zhao et al., 2022). How to prevent the neurodevelopmental deficiencies in these infants has been put in the center of child healthcare (Srinivas Jois, 2018). Current interventional strategies in promoting neurodevelopment among preterm infants are still lacking and less than optimal due to the underlying mechanisms of neurobehavioral development understudied in these high-risk population, which hindered the timely prevention, treatment, and prediction of neurobehavioral deficiencies in the early life stages.

The etiologies of preterm infant neurodevelopment are complex and multifactorial. We recently found that cumulative pain/stress experiences in early life are significantly related to altered neurobehavioral responses in preterm infants (Zhao et al., 2022), but the mechanisms demand further investigation. The brain-gut-microbiome axis, in which intestinal microbiome is proposed to play a key role in the regulation of stress and early programming of neuro-immune system that has been found to influence all aspects of human behaviors (Aatsinki et al., 2019; Oliphant et al., 2021; Seki et al., 2021). Preclinical and clinical studies have shown the brain-gut-microbiome axis involved in the regulation of neurobehavioral and cognitive development (Oliphant et al., 2021; Olson et al., 2021). Studies have reported that the gut microbiota regulated the pathophysiologic process of brain injury and neurological developments among preterm infants (Stewart et al., 2013; Seki et al., 2021; Beghetti et al., 2022). Several gut bacteria species have been identified to be involved in behavior mitigation and cognitive adjustment (Sordillo et al., 2019; Rozé et al., 2020).

Identifying potential pathogens and the pathogenesis process of gut microbiota involved in neurobehavioral development among preterm infants will facilitate the early relief and treatment of neurobehavioral deficiencies. Many are still unknown regarding the impact of early life experiences combined with gut microbiota on neurobehavioral development among preterm infants and few studies used longitudinal cohort design. Therefore, our study aimed to explore the longitudinal impact of gut microbiota and daily painful/stress experiences on the neurobehavioral development among preterm infants during their NICU hospitalization.

## 2. Methods

### 2.1 Design

A longitudinal cohort study was conducted at two NICUs in the Northeastern U.S. from January 2014 to August 2017. Preterm infants were followed from admission into the NICUs until their 28^th^ postnatal day or discharge from the NICUs. The study protocol was approved by the institutional review board of the study hospital and the affiliated institute. Written informed consent was obtained from parents of the preterm infants.

### 2.2 Inclusion and exclusion criteria

Preterm infants were included if they were: 1) 0 - 7 days old after birth; 2) born at 28 to 32 weeks of gestational age (28 0/7 to 32 6/7); 3) negative drug exposure history (no illicit drug use during pregnancy). Exclusion criteria included: 1) infant mothers were younger than 18 years old; 2) severe periventricular/ intraventricular hemorrhage (≥ Grade III); 3) other known congenital anomalies.

### 2.3 Measurements

#### 2.3.1 Demographic and clinical data collection

Demographic and clinical characteristics including sex, gestational age (GA), delivery type, birth weight and length were recorded by research nurses. The severity of illness of the infant was measured using the Score for Neonatal Acute Physiology – Perinatal Extension-II (SNAPPE-II) (Richardson et al., 2001). Daily antibiotic use, feeding types (mother’s breast milk, donor’s milk, and formula milk) and frequencies, and painful/stress experiences during the NICU hospitalization were recorded by research nurses.

#### 2.3.2 Assessment of daily painful/stress experiences

The Neonatal Infant Stressor Scale (NISS) was used to assess daily painful/stress experiences (47 acute and 23 chronic procedures or events) in early life, which was modified from the Australia version in our previous study based on the NICU practice in the U.S. (Zhao et al., 2022). The intensities of acute and chronic painful/stressful procedures were categorized into five domains (1= not painful/stressful; 2 = a little; 3 = moderate; 4 = very; and 5 = extremely painful/stressful). The detailed painful/stressful procedures and categories were list in Supplementary file 1. Trained research nurses extracted the NISS data from the infant electronic medical record and documented the data into the Research Electronic Data Capture (REDCap) system (Harris et al., 2019). Weighted frequencies (acute) and hours of procedures (chronic) were calculated by timing the counts and intensities of each procedure in each day of NICU stay to generate daily acute pain/stress scores and chronic pain/stress scores following our protocol (Zhao et al., 2022).

#### 2.3.3 Fecal sample and gut microbiota

Daily fecal samples were collected during diaper change depending on whether an infant had stool. Samples were processed following our previous protocol (Cong et al., 2016). The fecal sample DNA extraction and processing followed our previous methods and procedures (Cong et al., 2016; Chen et al., 2021). The raw sequence data were processed by the Mothur 1.42.3 pipeline(Schloss et al., 2009) following Mothur miseq process and the miseq bash (Supplementary file 2) (Chen et al., 2021). The operational taxonomic units (OTUs) were determined by clustering reads to the SILVA 119 16S reference dataset at a 97% identity, and then performing de novo OTU clustering on reads that failed to cluster to a reference (Edgar, 2018). Taxonomic annotation was also determined by the SILVA 119 V4 16□S rRNA reference database (Quast et al., 2013; Yilmaz et al., 2014).

#### 2.3.4 Neurobehavioral development assessment

Neurobehavioral outcomes were assessed using the NICU Network Neurobehavioral Scale (NNNS) (Lester et al., 2004) when an infant reached 36 to 38 weeks post-menstrual age (PMA) before the NICU discharge. The NNNS includes 115 items resulting in 13 summary scores assessing habituation, attention, arousal, self-regulation, handling, quality of movement, excitability, lethargy, reflexes, asymmetrical responses, hypertonicity, hypotonicity, and stress/abstinence. One trained and certified NNNS examiner who was blinded to all other assessments completed all the assessment and scoring of the NNNS subscales.

### 2.4 Data analysis

The demographic and clinical data, and OTU tables were imported into R 3.6.2 for statistical analysis. The clinical variables including the painful/stressful procedures of different levels and the population daily feeding of mother’s breast milk, donor’s milk and formula milk were visualized by plotting the pattern over time using the “ggplot2” package in R (Wickham, 2016). The sex differences regarding the demographic and clinical characteristics were tested by Wilcoxon rank sum test for continuous variable and Fisher’s exact test for categorical variables.

To explore the predictive microbiome biomarkers and estimate time-varying dynamics of their impact during early postnatal stage on the neurobehavioral outcomes of the preterm infants, sparse log-contrast regression with functional compositional predictors (Sun et al., 2020) was adopted. Infants who had 5 or more fecal samples after raw sequencing data processed were included in the current analysis to explore the time-varying effects using the sparse log-contrast regression model. The core OTUs were screened by the abundance and prevalence criteria before fitting the statistical model. Demographics variables including sex and race, delivery type, premature rupture of membranes (PROM) status and gestational age at birth were incorporated into the model as time-invariant control variables. The cubic spline basis was used for modeling the time-varying effects of the OTUs and a constrained group lasso (CGL) algorithm was used for compositional component selection at OTU level (Yuan and Lin, 2006; Sun et al., 2020). A hundred bootstrap samples were generated and used to provide supporting evidence for the stability of the results. The OTUs were chosen by the model selection process and those with higher proportions of being selected in the bootstrap procedure were kept.

## 3. Results

### 3.1 Demographic and clinical characteristics

A total of 92 preterm infants were recruited, and 55 infants were included in this report based on the completion of the microbiome and NNNS measurements (Supplementary Figure 1). The majority infants were Non-Hispanic/Latino White (74.55%), female (54.55%), and born through c-cession (61.82%) (Table 1). About 80% of the preterm infants received antibiotics during the first 3 days of the NICU stay; after day 3, only 20% of them used antibiotics. Feeding patterns included mother’s breast milk breastfeeding (61.65%), human donor milk (26.31%) and formula milk (12.04%) during the NICU hospitalization. The proportion of mother’s breast milk intake are shown in Supplementary Figure 2a, and sex-specific daily feeding patterns are shown in Supplementary Figure 3. For the daily average painful/stress experience (NISS scores), weighted frequencies of acute painful events (mean = 62.66, SD = 9.94) and weighted hours of chronic painful procedures (mean = 89.84, SD = 36.72) were calculated and plotted across sex (Supplementary Figure 2b, 2c).

**Table 1.** Characteristics of the included infants, Mean (SD)

|  | Total (n = 55) | Female (n=30) | Male (n=25) |
| --- | --- | --- | --- |
| Birth gestational age (week) | 30.72 (1.71) | 30.53 (1.72) | 30.96 (1.71) |
| Birth body length (cm) | 39.94 (3.25) | 39.56 (3.34) | 40.38 (3.17) |
| Birth body weight (g) | 1444.53 (406.61) | 1362.87 (413.29) | 1542.52 (383.76) |
| Birth head circumference (cm) | 27.86 (1.88) | 27.50 (2.05) | 28.31 (1.59) |
| SNAPE II [media, IQR] <sup>a</sup> | 9.31 (9.66) | 11.07 (10.57) | 7.2 (8.15) |
| C-cession (n, %) | 34 (61.82%) | 21 (70.00%) | 13 (52.00%) |
| Pre-rupture of membrane (n, %) | 22 (40%) | 11 (36.67%) | 11 (44%) |
| Race (n, %) |  |  |  |
| White | 41 (74.54%) | 23 (76.67%) | 18 (72.00%) |
| American African | 11 (20.00%) | 4 (13.33%) | 7 (28.00%) |
| Asian | 2 (3.64%) | 2 (6.67%) | 0 (0.00%) |
| Unknown | 1 (1.82%) | 1 (3.33%) | 0 (0.00%) |
| Averaged MBM percentage <sup>b</sup> | 61.46% (32.07) | 63.09% (33.02) | 59.51% (31.46) |
| Averaged Acute pain <sup>c</sup> | 62.66 (9.94) | 62.01 (9.65) | 63.43 (10.42) |
| Averaged Chronic pain <sup>c</sup> | 89.84 (36.72) | 93.44 (37.76) | 85.82 (35.68) |

<sup>a</sup> SNAPE II, Score for Neonatal Acute Physiology – Perinatal Extension-II (SNAPPE-II)
<sup>b</sup> MBM, Mother's Breast Milk
<sup>c</sup> Weighted frequencies (acute) and hours of procedures (chronic) were calculated by timing the counts and intensities of each procedure in each day of NICU stay to generate daily acute pain/stress scores and chronic pain/stress scores.

### 3.2 The gut microbiota compositions

A total of 584 stool samples were included in the analysis (Supplementary Table 1). The most abundant phylum was *Proteobacteria, Firmicutes*, and *Bacteroidetes*. The compositional relative abundances for the 55 preterm infants were plotted on the average basis (Figure 1). Detailed taxonomy of each OTU was summarized in Supplementary Table 2.

**Figure 1.**
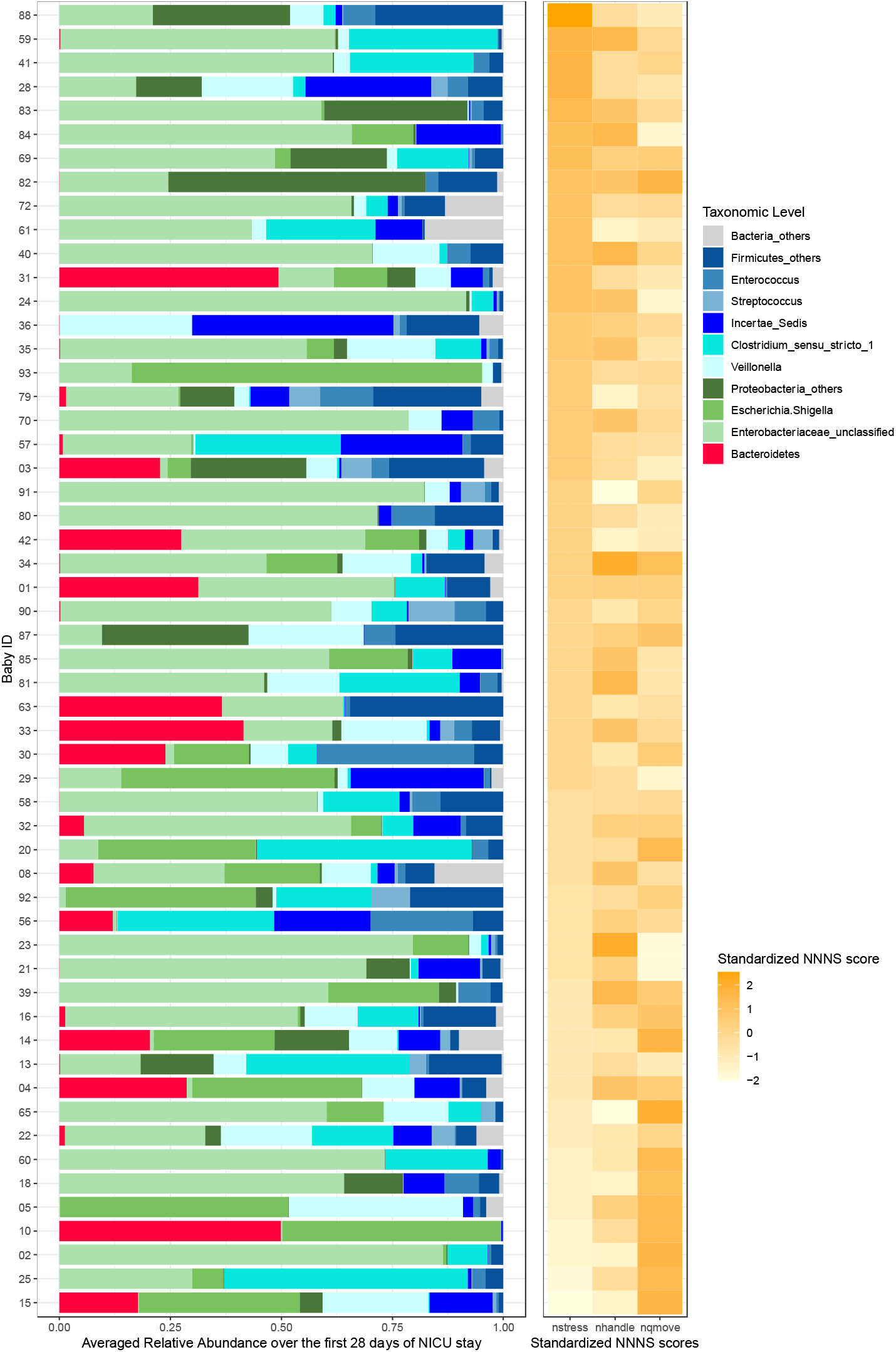
Relative abundance of gut microbiota and 3 sub-scales for each infant a. Relative abundance of gut microbiota for each infant b. Standardized scores of NNNS subscales for each infant The infants were ordered according to the standardized score of NSTRESS scores. A standardized score of NNNS subscales (NSTRESS, NQMOVE, and NHANDLING) was generated by dividing the difference between each infant’s score and the mean by the standard deviation. The standardized scores of each these subscales were plotted with the gut microbiome for each infant.

### 3.3 Neurobehavioral development

The NNNS assessment scores were presented in Table 2 and Supplementary Figure 4. These preterm infants had high level of hypertonicity, hypotonicity, and asymmetric reflexes (median score = 0), followed by stress/abstinence and handling (Supplementary Figure 4). Given the substantial missing values on some of the subscales, the main focuses of the current analysis were on stress/abstinence (NSTRESS), handling (NHANDLING), and quality of move (NQMOVE). The stress/abstinence (NSTRESS), handling (NHANDLING), and quality of move (NQMOVE) among these preterm infants at 36 to 38 weeks of post-menstrual age was 0.18 (SD = 0.09), 0.56 (SD = 0.21), and 3.97 (SD = 0.62), respectively. There was no significant difference between females and males.

**Table 2.** Neurobehavioral outcomes of the included infants, Mean (SD)

| NNNS <sup>a</sup> | Total (n = 55) | Female (n=30) | Male (n=25) |
| --- | --- | --- | --- |
| Stress/abstinence (NSTRESS) | 0.18 (0.09) | 0.19 (0.09) | 0.17 (0.09) |
| Handling (NHANDLING) | 0.56 (0.21) | 0.57 (0.21) | 0.56 (0.22) |
| Quality of movement (NQMOVE) | 3.97 (0.62) | 3.90 (0.65) | 4.05 (0.60) |

<sup>a</sup> NNNS, NICU Network Neurobehavioral Scale

### 3.4 Associations of pain/stress experience and gut microbiota with neurobehaviors

The estimated coefficients of the control variables for the NSTRESS, NQMOVE, and NHANDLING assessment were shown in Supplementary Table 3. As shown by the bootstrap analysis, infants with older birth GA, sex of male, race of white, virginal birth, no RPOM, lower acute pain, higher kangaroo care, and no antibiotic uses in the first 3 days of NICU stay might be associated with better outcomes including lower NSTRESS scores (Figure 2 a) and lower NHANDLING scores (Figure 3 a), and higher NQMOVE scores (Figure 4 a). In particular, the positive association of higher acute pain/stress (NISS score) with higher NSTRESS scores was seen in close to 95% of bootstrap (Figure 2 a), indicating that infants who experienced less acute painful/stressful events during the NICU stay had better neurobehavioral outcomes. However, the relationships of feeding patterns and chronic pain (NISS score) with NNNS subscales were still undetermined.

**Figure 2.**
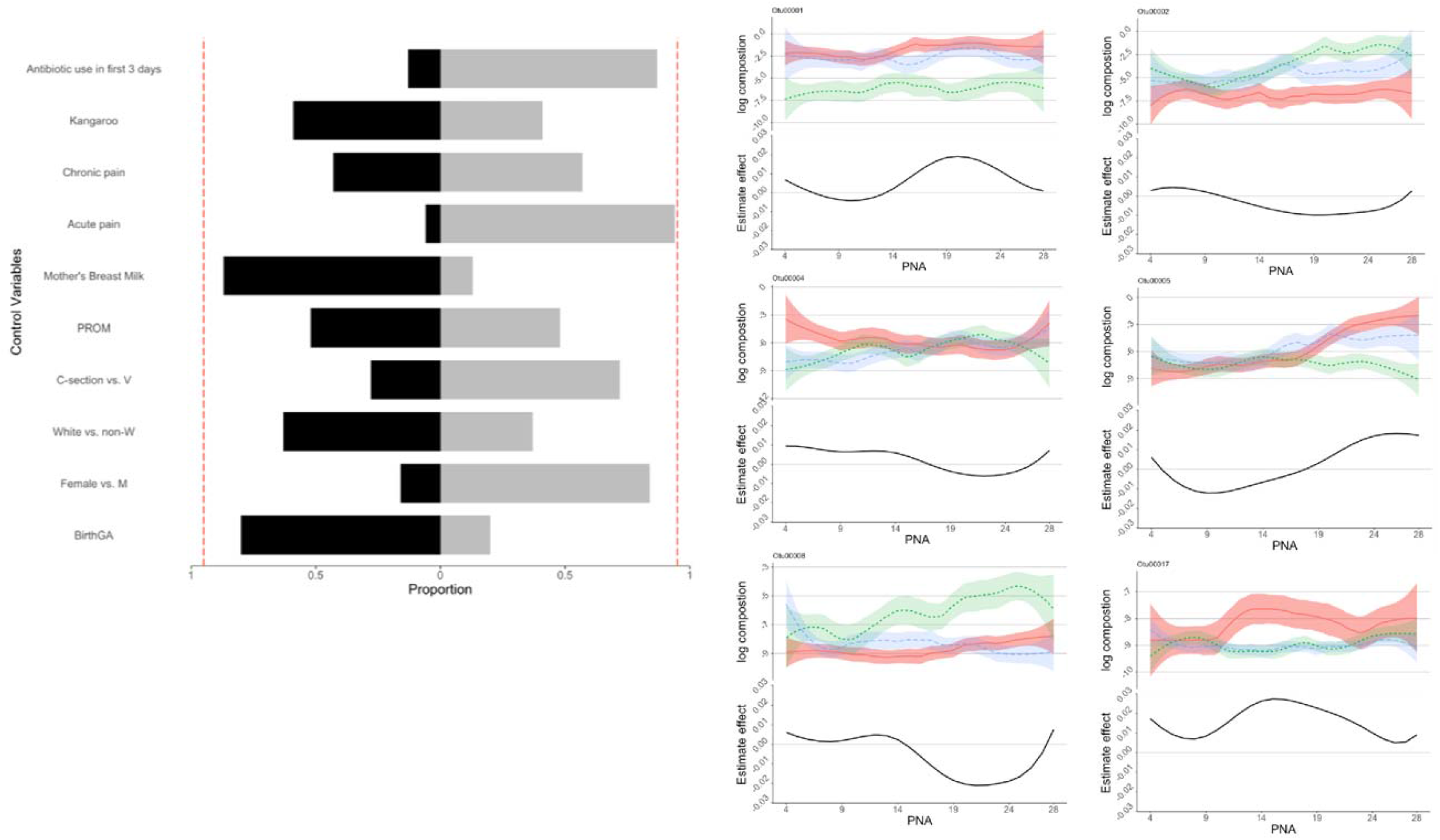
Control sign and OTUs for NSTRESS a. Control sign and NSTRESS b. OTUs associated with NSTRESS scores Figure 2 a, the proportions of the signs of the estimated coefficients of the control variables. Proportions of negative signs were shown as black blocks to the right, and those of positive signs were shown as light gray blocks to the left. Lower NSTRESS scores indicated better development, those variables in black indicated association with lower NSTRESS scores. The red dotted lines show the 90% of bootstrap. Figure 2 b1-6, the estimated time-varying effects of OUT on NSTRESS scores during the NICU stay.

**Figure 3.**
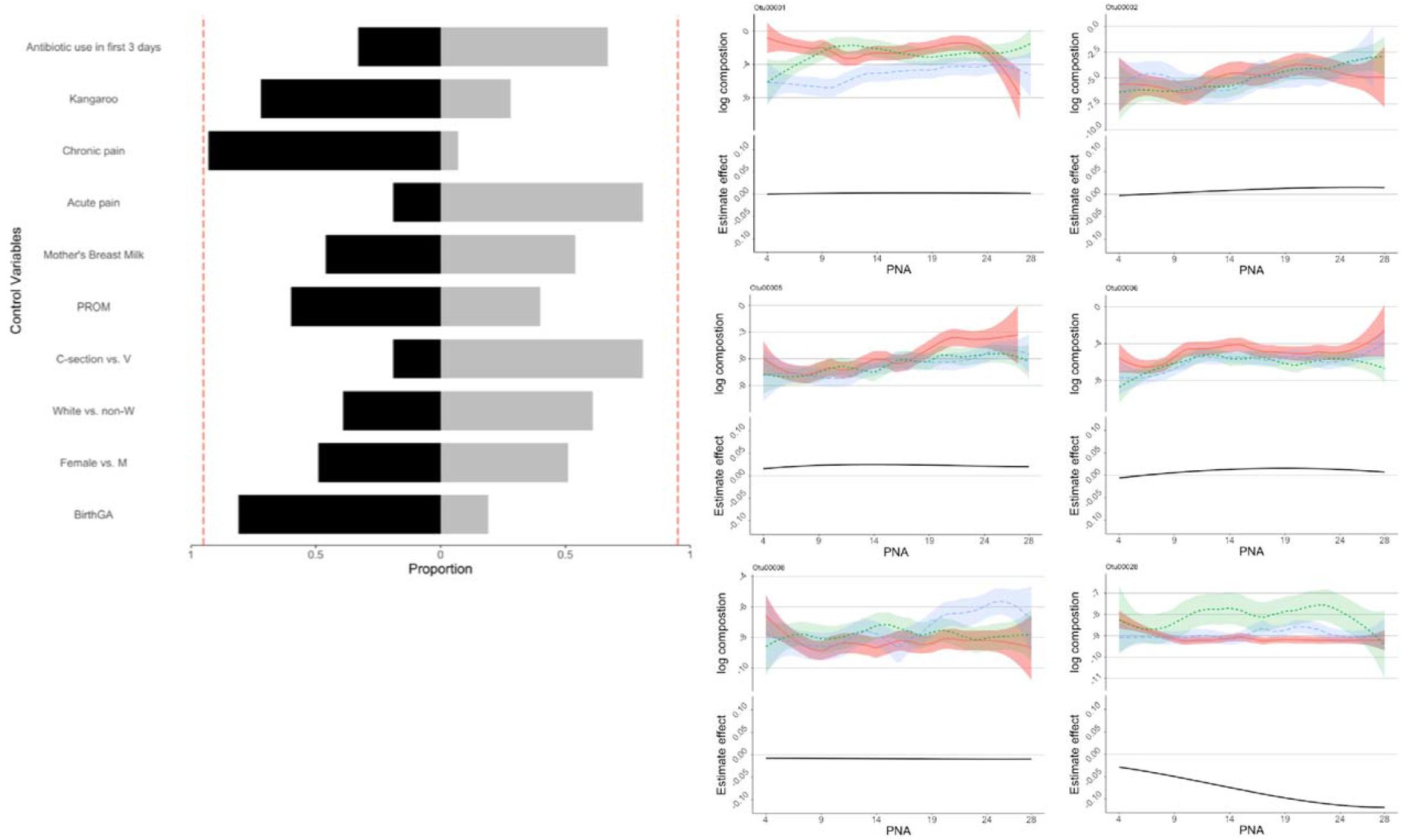
Control sign and OTUs for NHANDLING a. Control sign and NHANDLING b. OTUs associated with NHANDLING scores Figure 3 a, the proportions of the signs of the estimated coefficients of the control variables. Proportions of negative signs were shown as black blocks to the right, and those of positive signs were shown as light gray blocks to the left. Lower NHANDLING scores indicated better development, those variables in black indicated association with lower NHANDLING scores. The red dotted lines show the 90% of bootstrap. Figure 3 b1-6, the estimated time-varying effects of OUT on NSTRESS scores during the NICU stay.

**Figure 4.**
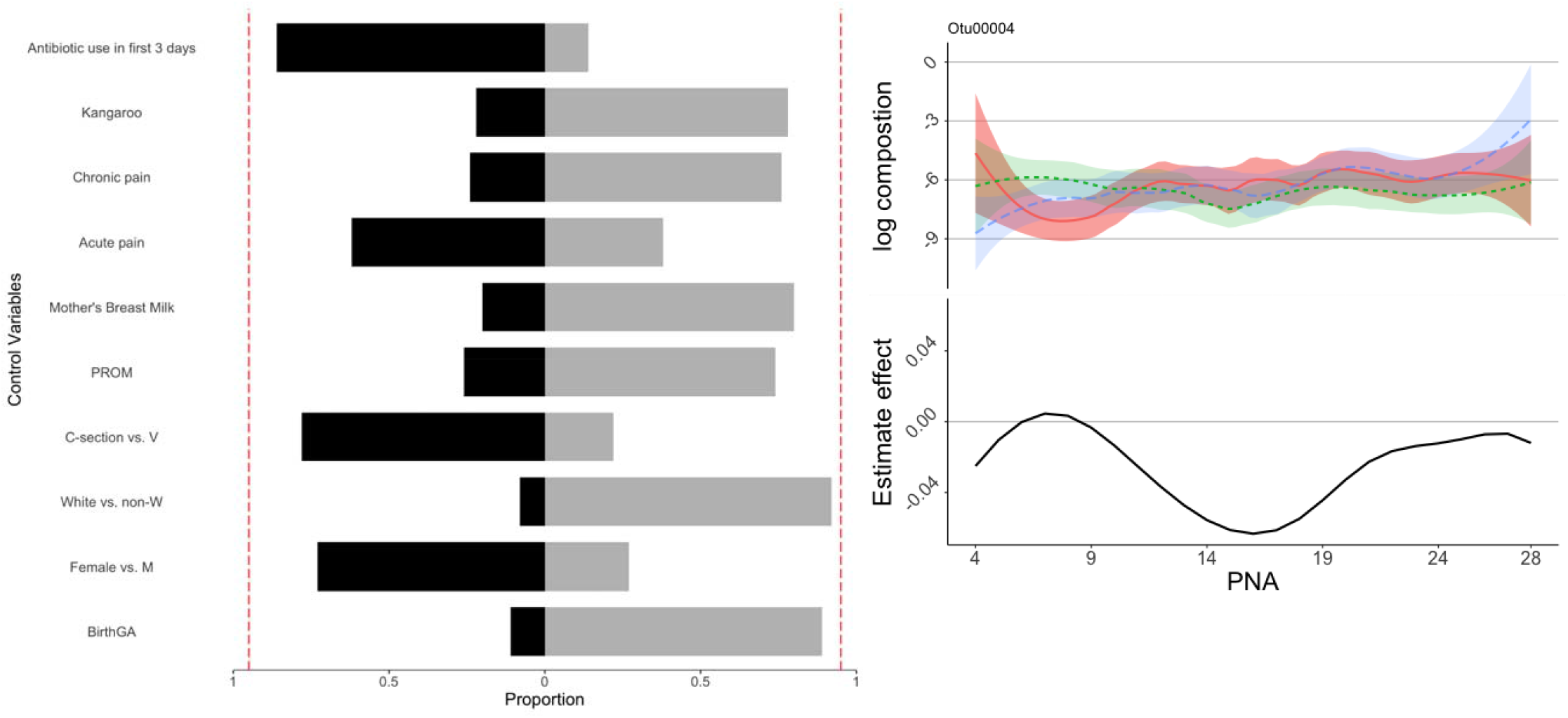
Control sign and OTUs for NQMOVE a. Control sign and NQMOVE b. OTUs associated with NQMOVE scores Figure 4 a, the proportions of the signs of the estimated coefficients of the control variables. Proportions of negative signs were shown as black blocks to the right, and those of positive signs were shown as light gray blocks to the left. Higher NQMOVE scores indicated better development, those variables in light gray indicated association with higher NQMOVE scores. The red dotted lines show the 90% of bootstrap. Figure 4 b, the estimated time-varying effects of OUT on NQMOVE scores during the NICU stay.

To illustrate the relationships between gut microbiota compositions and NNNS subscales (NSTRESS, NHANDLING, and NQMOVE), the standardized scores of each these subscales were plotted with the gut microbiome for each infant using a heatmap (Figure 1). A standardized score of NNNS subscales was generated by dividing the difference between each infant’s score and the mean by the standard deviation.

Eight OTUs were identified to be associated with NNNS subscales through the regression analysis (Figure 2, 3, and 4). At the taxonomy levels, five belong to *Firmicutes* (OTU4, OTU5, OTU6, OTU8, and OTU28), and three belong to *Proteobacteria* (OTU1, OTU2, and OTU17). The taxa of the OTU1 (*Enterobacteriaceae_unclassified*), OTU2 (*Escherichia-Shigella*), and OTU17 (*Enterobacteriaceae_unclassified*) were identical at the family level (*Enterobacteriaceae*), OTU1 (*Enterobacteriaceae_unclassified*) and OTU17 (*Enterobacteriaceae_unclassified*) were also identical at genus level (Supplementary Table 2). The associations of these OTUs and NNNS subscales varied depending on different days of NICU stay (Figure 2, 3, and 4) after controlling for feeding types and pain/stress experiences in addition to demographic and clinical characteristics.

The OTU1 (*Enterobacteriaceae_unclassified*), OTU2 (*Escherichia-Shigella*), OTU5 (*Veillonella*), OTU4 (*Incertae_Sedis*), OTU8 (*Clostridium_sensu_stricto_1*), and OTU17 (*Enterobacteriaceae_unclassified*) were identified to be associated with NSTRESS, and their estimated time-varying effects are presented in each sub figures of Figure 2. The effect of the OTU2 (*Escherichia-Shigella*), OTU4 (*Incertae_Sedis*), and OTU8 (*Clostridium_sensu_stricto_1*) on the NSTRESS score switches from positive to negative during the postnatal days from 4 to 28, while the effect of the OTU1 (*Enterobacteriaceae_unclassified*) and OTU5 (*Veillonella*) switches from negative to positive. The OTU17 (*Enterobacteriaceae_unclassified*) shows consistently positive effect on NSTRESS score during the 28 days of NICU stay. Elevated abundance of *Enterobacteriaceae* (OTU1 and OTU17) was significantly associated with increased NSTRESS, particularly after two weeks of NICU stay (Figure 2). But the elevated enrichment of *Escherichia-Shigella* (OTU2) was associated with decreased NSTRESS, particularly after 10 days.

The OTU1 (*Enterobacteriaceae_unclassified*), OTU2 (*Escherichia-Shigella*), OTU 5 (*Veillonella*), OTU 6 (*Enterococcus*), OTU8 (*Clostridium_sensu_stricto_1*), and OTU28 (*Streptococcus*) were selected for the model regressing on NHANDLING. Their estimated time-varying effects are presented in each sub figures of Figure 3. The effect of the OTU1 (*Enterobacteriaceae_unclassified*), OTU2 (*Escherichia-Shigella*), OTU5 (*Veillonella*), and OTU6 (*Enterococcus*) on the NHANDLING score remains positive during the first month, while the effect of the OTU8 (*Clostridium_sensu_stricto_1*) was constantly negative. The OTU28 (*Streptococcus*) shows enlarging negative effect on the NHANDLING score over the first month.

The OTU4 (*Incertae_Sedis*) was the only OTU selected for NQMOVE; their estimated time-varying effects are presented in Figure 4. The effect of the OTU4 (*Incertae_Sedis*) on the NQMOVE score became negative after day 10.

## 4. Discussion

Our study using a longitudinal modeling approach demonstrated the impact of early life pain/stress experience and gut microbiota on neurobehavioral outcomes in preterm infants during their NICU hospitalization. Consistent with previous studies, our findings showed that preterm infants had a higher risk of neurobehavioral deficiency compared with full term infants (Kiblawi et al., 2014; Provenzi et al., 2018; McGowan et al., 2022). In comparison to the neurobehavioral results from healthy full-term infants at birth (Provenzi et al., 2018), our findings showed that preterm infants had higher NNNS scores than full term infants in stress/abstinence (0.18 vs. 0.11) and handling responses (0.56 vs. 0.38), and lower quality of movement (3.97 vs. 4.71). The negative impact of higher acute painful/stressful events during the NICU stay on worse neurobehavioral outcomes is congruent with previous studies (Swamy et al., 2008; Lavanga et al., 2021; Russell et al., 2021). We identified eight OTUs of gut microbiome that were significantly associated with infant neurobehavioral profiles in early life. Most importantly, our study uncovered potential pathogenesis process of *Enterobacteriaceae* and *Streptococcaceae* involved in neurobehavioral outcomes by depicting the dynamical impacts of OTUs on NNNS scores. These findings are consistent with previous studies, which showed the brain-gut-microbiome axis involved in neonatal brain damage and immunity (Currie et al., 2011; Seki et al., 2021), and influencing the lifelong health of humans (Lyte, 2014; Cong et al., 2015).

The role of *Enterobacteriaceae* on NSTRESS is still unclear given that elevated abundance of *Enterobacteriaceae unclassified* (OTU1 and OTU17) was significantly associated with increased NSTRESS, but the elevated enrichment of *Escherichia-Shigella* (OTU2) was associated with decreased NSTRESS (Figure 2). Enriched *Enterobacteriaceae* has been demonstrated to induce inflammatory and stress response (Kim et al., 2017). Some studies reported the harmful effect of *Enterobacteriaceae* on cognitive function (Wasser et al., 2020; Streit et al., 2021), but the role of *Escherichia-Shigella* is unclear. Of note, only 55% percent of OTU2 was *Escherichia-Shigella*, the other 45% is unknown (Supplementary Table 4). Our study also found negative association between enriched abundance of *Incertae_Sedis* (OTU4) and *Veillonella* (OTU5) and lower abstinence/stress level (NSTRESS) after 14 days, which may indicate the neuro-protective effect of *Incertae_Sedis* and *Veillonella*. These potential protective effects were supported by previous studies which reported the role that *Incertae_Sedis* plays in allergic disease (Quast et al., 2013), and *Veillonella* played in energy conservation among infants (Wang et al., 2020). The role of *Incertae_Sedis* and *Veillonella* in the first two weeks of NICU hospitalization need more investigations.

Our study found that enrichment of *Enterococcus* (OTU6), genus level of *Enterococcaceae*, was associated with better handling response (Figure 3). The protective role of *Enterococcaceae* in the gut among cancer patients receiving radiotherapy was reported in a previous study that it is involved in maintaining hematopoiesis and intestinal barriers (Guo et al., 2020). The elevated abundance of *Enterococcaceae* has been reported to exert in the pathophysiology progress of several disorders such as infection and cytokines response (Hu et al., 2019; Rabe et al., 2020). The negative effect of *Clostridium_sensu_stricto_1*(OTU8) on NHANDLING was also confirmed in our study, since it was associated with higher risk of necrotizing enterocolitis and prematurity (Schönherr-Hellec et al., 2018).

Enriched *Streptococcus* (OTU28) was related to a lower score of NHANDLING indicating a better developmental outcome, and the negative impact accumulated over time in NICU. Aatsinki et al reported the positive association between behavioral development and *Streptococcus* in infants at the age of 6 months (Aatsinki et al., 2019), but the role of enriched *Streptococcus* in preterm infants is still unknown. Elevated *Streptococcus* (OTU28) was found in the gut microbiota among children in atopic dermatitis, an allergic reaction (Park et al., 2020). A previous study also reported the role of *Streptococcus* (OTU28) in infections, i.e., sepsis and meningitis among preterm infants (Geetha et al., 2021) and *Streptococcus* pneumoniae in infants (Turner et al., 2012; Nieto-Moro et al., 2020), a possible reason might be related to the immature immune responses (Currie et al., 2011).

Even the negative effect of *Clostridium_sensu_stricto* (OTU8) *and Streptococcus* (OTU28) on handling response was identified (Figure 3), our study did not find significantly direct association between breastfeeding and neurobehavioral development. Higher portion of breastfeeding could alter gut microbiota compositions (Pannaraj et al., 2017), as well as was associated with better neurobehavioral outcomes (Zhao et al., 2022). However, previous studies reported inconsistent findings regarding the effect of breastfeeding on *Clostridium_sensu_stricto and Streptococcus*. One study reported that higher proportion of breastfeeding and human donor milk could significantly increase the enrichment of *Clostridium_sensu_stricto* among preterm infants (Aguilar-Lopez et al., 2021). Another study also reported the protection role of breastfeeding on decreasing the risk of *Streptococcus* induced infection (Béghin et al., 2021). Further studies are needed to uncover the entangling between breastfeeding, gut microbiota, and neurobehavior development among preterm infants.

The sex and race differences of neurobehavioral development in preterm infants warrant more effort to investigate the possible underneath mechanisms, even there was no significant findings from the current study. Evidence has confirmed that there exists an impact of sex-dependent gut microbiota on the behavioral development of full-term infants (Jiang et al., 2019; Chen et al., 2021; Laue et al., 2022). The gut microbiota compositions and predicted functions differences between females and males could be a possible reason (Cong et al., 2016; Chen et al., 2021, 2022). Previous studies also reported the race differences of gut microbiota diversities (Pannaraj et al., 2017; Stearns et al., 2017), and compositions (Stearns et al., 2017; Cheng et al., 2022). Future studies should continue to investigate the mechanisms of sex and race disparities of neurobehavioral outcomes among preterm infants.

Our findings provided new evidence to demonstrate the gradually mature brain-gut-microbiome axis contributing to the neurobehavioral development among preterm infants. Manipulating the identified gut microbiota by interventional strategies such as fecal microbiome transplantation and/or supplementing prebiotics and probiotics may effectively improve the measured neurobehavioral outcomes among preterm infants, i.e., stress, handling responses and quality of movement (Cryan and Dinan, 2012; Sarkar et al., 2018; Vogel et al., 2020; Samara et al., 2022). The OTUs associated with neurobehavioral development among preterm infants identified in the current study were generated using the 16S RNA sequencing data that may have limitations in conducting data analysis and making inferences based on OTUs, i.e., less powerful to detect differential effects and functions of gut microbiota. Further studies may also need to employ shotgun sequencing and brain image techniques to yield more information including the metabolic functions of the gut microbiome community and the activities of the brain-gut-microbiome axis to explore how the gut microbiome and host brain-gut axis function in the growth and development among preterm infants.

## 5. Strengths and Limitations

To the best of our knowledge, this is one of the first longitudinal studies modeling the impact of early life pain/stress experience and gut microbiota on neurobehavioral outcomes among preterm infants throughout the NICU hospitalization. The neurobehavioral development measured by NNNS in this study may serve as valid indicators to predict neurodevelopmental and infant health outcomes in the clinical settings, although it may not directly predict infant mortality or morbidity. One of the limitations of our study was that this study only included preterm infants born at 28 to 32 weeks of gestational age which did not consider the extremely preterm infants who are more likely to have developmental deficits. Another limitation was the weakness of the 16S sequencing data and analysis pipeline based on OTUs. Therefore, the generalization of findings from this study should be cautious and applying evidence generated in this study should be prudent.

## 6. Conclusions

This longitudinal cohort study reported the dynamic impact of gut microbiota and adverse life experiences in the NICU on neurobehavioral development among preterm infants. In addition, specific OTUs combined with acute painful/stressful experiences were determined to influence infants’ neurobehavioral development during their NICU hospitalization. Further studies may need to develop interventions targeting these factors to prompt developmental outcomes in this population.

## Data Availability

The raw sequence data were achieved in NCBI (https://submit.ncbi.nlm.nih.gov/subs/sra/SUB8904718/). Deidentified data will be available upon reasonable request. Requests to access these datasets should be directed to.

## Ethics Statement

The studies involving human participants were reviewed and approved by institutional review board in the University of Connecticut and Connecticut Children’s Medical Center. The deidentified dataset was used in the analysis. Written informed consent to participate in this study was provided by the participants’ legal guardian/next of kin.

## Author Contributions

Conceptualization, J.C., X.C., T. Z., and K.C.; formal analysis, J.C., H.L., T.Z., Z.S., W.X., K.C., M.H.C., and X.C.; funding acquisition, X.C.; methodology, J.C., H.L., W.X., and X.C.; project administration, W.X., and X.C.; writing-original draft, J.C., H.L., T.Z., and X.C.; writing-review and editing, All. All authors critically revised the manuscript, gave final approval, and agree to be accountable for all aspects of work ensuring integrity and accuracy.

## Acknowledgments

The authors would like to acknowledge all the infants and their parents’ participants in this study. The authors thank the medical and nursing staff in the NICUs of Connecticut Children’s Medical Center at Hartford and Farmington, CT for their support and assistance. The authors thank Victoria Vazquez, Shari Galvin, and Megan Fitzsimons for their assistance in recruiting subjects, collecting clinical data, and managing stool sample collection and storage. The authors would also like to acknowledge the support from the Bio-Behavioral Lab (BBL) in the University of Connecticut School of Nursing. The authors acknowledge the Microbial Analysis, Resources, and Services (MARS) facility at the University of Connecticut for their ongoing support of this project.

## Funding Resource

This publication was supported by the National Institute of Nursing Research of the National Institutes of Health (NIH-NINR) under Award Number K23NR014674 and R01NR016928, and Affinity Research Collaboratives award through the University of Connecticut Institute for Systems Genomics. Jie Chen received research support from Virginia Stone Fund through American Nurses Foundation Research Grants Award, Eastern Nursing Research Society (ENRS)/Council for the Advancement of Nursing Science Dissertation Award, Sigma Theta Tau International Mu Chapter Research Award, and the University of Connecticut Dissertation Fellowship.

