## Supplementary material for "The impact of early life experiences and gut microbiota on neurobehavioral development among preterm infants: A longitudinal cohort study": S Figure 1 CONSORT Form.docx

Preterm infant recruited (n=92)

Infant had fecal samples and NNNS assessment (n=87)

- 5 infants had no fecal sample left after Mothur analysis

Infant had fecal samples and NNNS assessment (n=82)

Infant included in final analysis

(n=55)

- 1 infant did not have fecal sample and NNNS assessment
- 4 infants did not have NNNS assessment
- 27 infants had 4 or less fecal sample left after Mothur analysis

S Figure 1 CONSORT Form
