## Supplementary material for "The impact of early life experiences and gut microbiota on neurobehavioral development among preterm infants: A longitudinal cohort study": S Table 3 Estimation of Control Variables.pdf

| | GA | Female vs. Male | White vs. Non-White | C-section vs. Vaginal | PROM vs. N-PROM | MBM proportion | Acute pain/stress | Chronic pain/stress | kangaroo care | Antibiotics use in the first 3 days | Intercept |
|---|---|---|---|---|---|---|---|---|---|---|---|
| NSTRESS | -0.012 | 0.019 | -0.007 | 0.018 | -0.007 | -0.061 | 0.002 | 0.001 | -0.001 | 0.048 | 0.495 |
| NHANDLING | -0.043 | 0.007 | -0.041 | 0.103 | -0.05 | 0.001 | 0.008 | -0.029 | -0.001 | 0.088 | 1.294 |
| NQMOVE | 0.128 | -0.112 | 0.389 | -0.199 | 0.148 | 0.268 | -0.004 | 0.024 | 0.007 | -0.314 | -0.455 |

Note: PROM, Pre-rupture of membrane; MBM, Mother's Breast Milk.
