## Supplementary material for "The impact of early life experiences and gut microbiota on neurobehavioral development among preterm infants: A longitudinal cohort study": S Table 4 Selected 8 OTUs associated with of NNNS subscales.pdf

| NSTRESS | NHANDLING | NQMOVE | OTU | Size | Taxonomy |
|---|---|---|---|---|---|
| Selected | Selected | NA | Otu00001 | 13859029 | Bacteria(100);Proteobacteria(100);Gammaproteobacteria(100);Enterobacteriales(100); Enterobacteriaceae(100);Enterobacteriaceae_unclassified(95); |
| Selected | Selected | NA | Otu00002 | 9732098 | Bacteria(100);Proteobacteria(100);Gammaproteobacteria(100);Enterobacteriales(100); Enterobacteriaceae(100);Escherichia-Shigella(55); |
| Selected | NA | Selected | Otu00004 | 2880234 | Bacteria(100);Firmicutes(100);Clostridia(100);Clostridiales(100); Peptostreptococcaceae(100);Incertae_Sedis(100); |
| Selected | Selected | NA | Otu00005 | 2667454 | Bacteria(100);Firmicutes(100);Negativicutes(100);Selenomonadales(100); Veillonellaceae(100);Veillonella(100); |
| NA | Selected | NA | Otu00006 | 2390001 | Bacteria(100);Firmicutes(100);Bacilli(100);Lactobacillales(100); Enterococcaceae(99);Enterococcus(99); |
| Selected | Selected | NA | Otu00008 | 1576632 | Bacteria(100);Firmicutes(100);Clostridia(100);Clostridiales(100); Clostridiaceae_1(100);Clostridium_sensu_stricto_1(100); |
| Selected | NA | NA | Otu00017 | 168136 | Bacteria(100);Proteobacteria(100);Gammaproteobacteria(100);Enterobacteriales(100); Enterobacteriaceae(100);Enterobacteriaceae_unclassified(100); |
| NA | Selected | NA | Otu00028 | 92536 | Bacteria(100);Firmicutes(100);Bacilli(100);Lactobacillales(100); Streptococcaceae(100);Streptococcus(100); |

Note: NA, not applicable.
