## Supplementary material for "The impact of early life experiences and gut microbiota on neurobehavioral development among preterm infants: A longitudinal cohort study": Supplementary file 1.docx

**Coding system for events of different levels of pain/stress in NISS assessment**

The Neonatal Infant Stressor Scale (NISS) was used to assess daily painful/stress experiences (47 acute and 23 chronic procedures or events) in early life, which was modified from the Australia version in our previous study based on the NICU practice in the U.S. (Zhao et al., 2022).

Levels for pain/stress severity: Level 2=a little; Level 3=moderate; Level 4= very; Level 5=extremely.

Acute pain/stress events include level 2 (mouth care, learn bottle, nasal suction, learn breastfeeding); level 3 (position change, diaper change, continuous positive airway pressure manipulation, remove infant from incubator wrapped/unwrapped, being weighted, tape removal); level 4 (heel stick and suctioning); level 5 (intubation and multiple intravenous infusion).

Chronic pain/stress events include level 2 (nasogastric and orogastric tube in situ, intravenous in situ, peripherally inserted central catheter line in situ, umbilical venous and arterial catheter in situ, high-flow nasal cannula oxygen, phototherapy, replogle tube in situ, intranasal and head box oxygen); level 3 (continuous positive airway pressure manipulation, nothing by mouth, conventional ventilation with/without sedation, high-flow humidified nasal cannula oxygen, local infection, recovery from surgery, high-frequency oscillatory/Jet ventilation with sedation, chest tube placed on water seal); level 4 (having confirmed systemic infection, high-frequency oscillatory/Jet ventilation without sedation, and chest tube placed on wall suction).
