## Supplementary material for "The impact of early life experiences and gut microbiota on neurobehavioral development among preterm infants: A longitudinal cohort study": Supplementary file 2.docx

**Raw sequence data processing**

The raw sequence data were processed following MARS Kendra’s mothur batch file.

https://github.com/krmaas/bioinformatics/blob/master/mothur.batch

Bioinformatics Pipeline: mothur

The raw sequence data were achieved in NCBI (https://submit.ncbi.nlm.nih.gov/subs/sra/SUB8904718/).
