## Supplementary material for "The impact of early life experiences and gut microbiota on neurobehavioral development among preterm infants: A longitudinal cohort study": Supplementary files Legends 12022022.docx

Supplementary file 1

Coding system for events of different levels of pain/stress in NISS assessment

Supplementary file 2

Raw sequence data processing bash

S. Fig 1

CONSORT chart of the included infants

S. Fig 2

Feeding and NISS

a. Average proportion of mother’s breast milk intake feeding between females and males

b. Daily average weighted frequencies of acute pain/stress events between females and males

c. Daily average weighted hours of chronic pain/stress events between females and males

S. Fig 3

Daily feeding patterns for females and males

S. Fig 4

Neurobehavioral development

S. Table 1

Daily sample collection of each infant

S. Table 2

Taxonomy of each OTU

S. Table 3

Estimation of Control Variables: Results of regression models

S. Table 4

Selected 8 OTUs associated with of NNNS subscales
