## Supplementary figures and images for "The impact of early life experiences and gut microbiota on neurobehavioral development among preterm infants: A longitudinal cohort study"

### S Figure 2.pdf

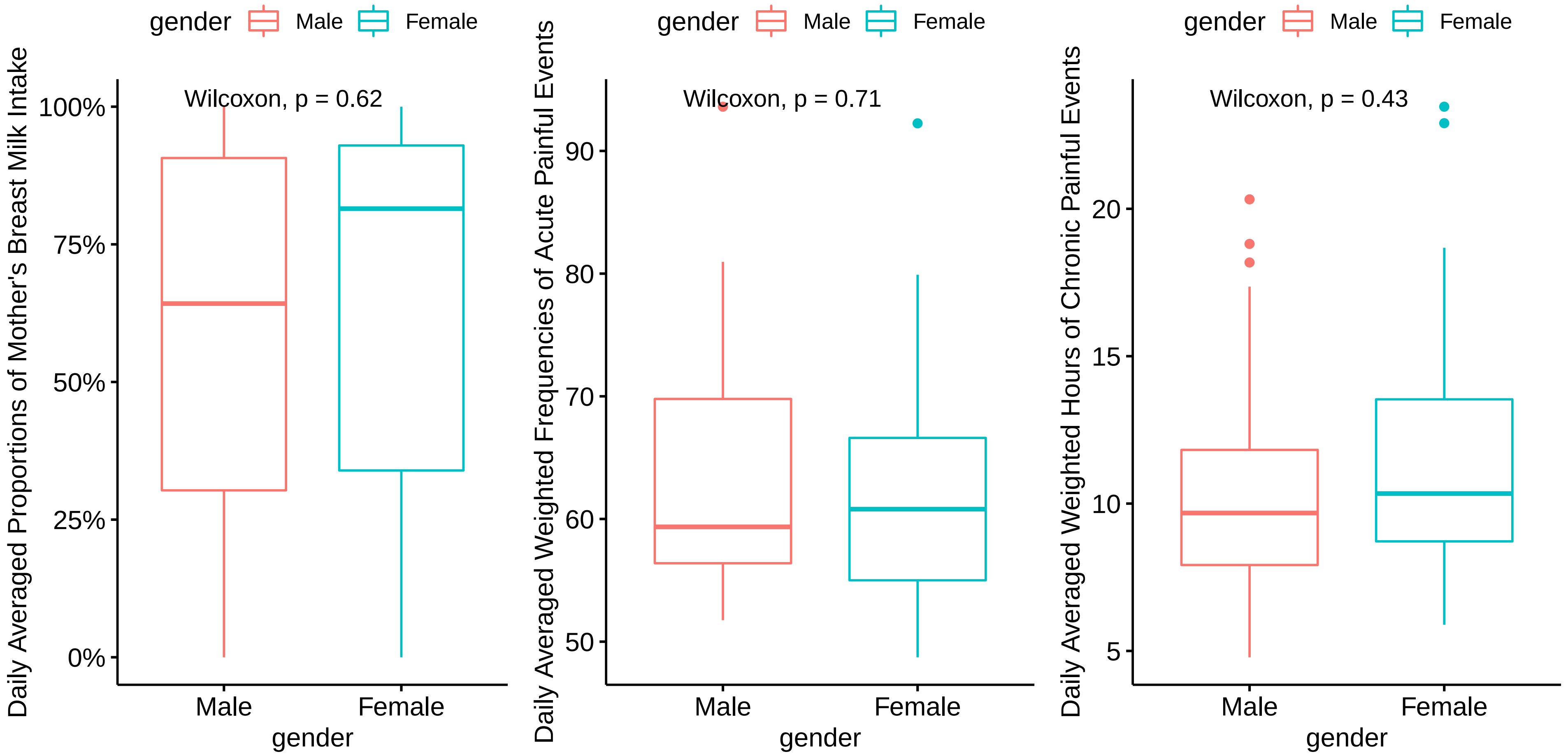

### S Figure 3 feed.pna.pdf

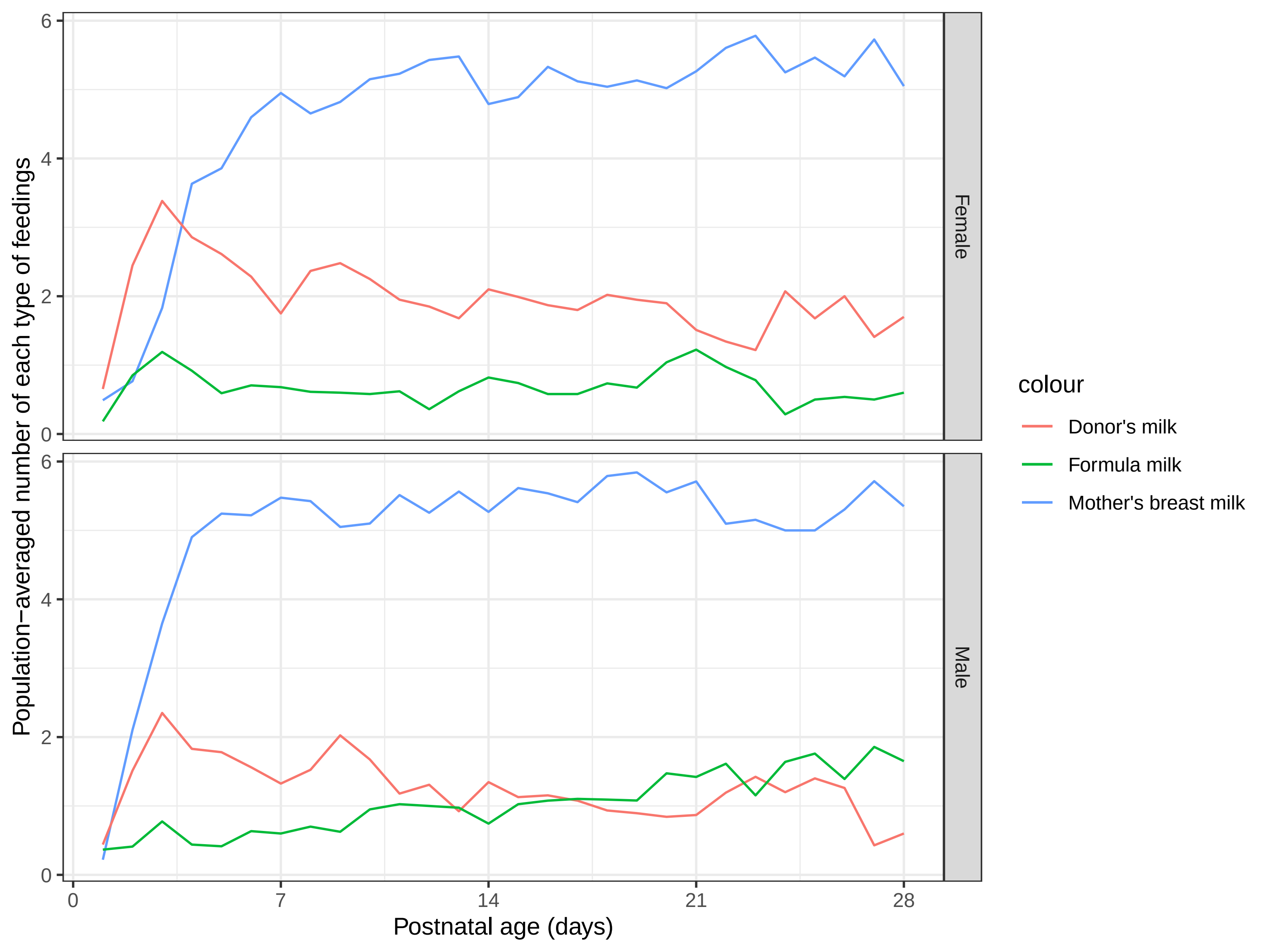

### S Figure 4 nnns.pdf

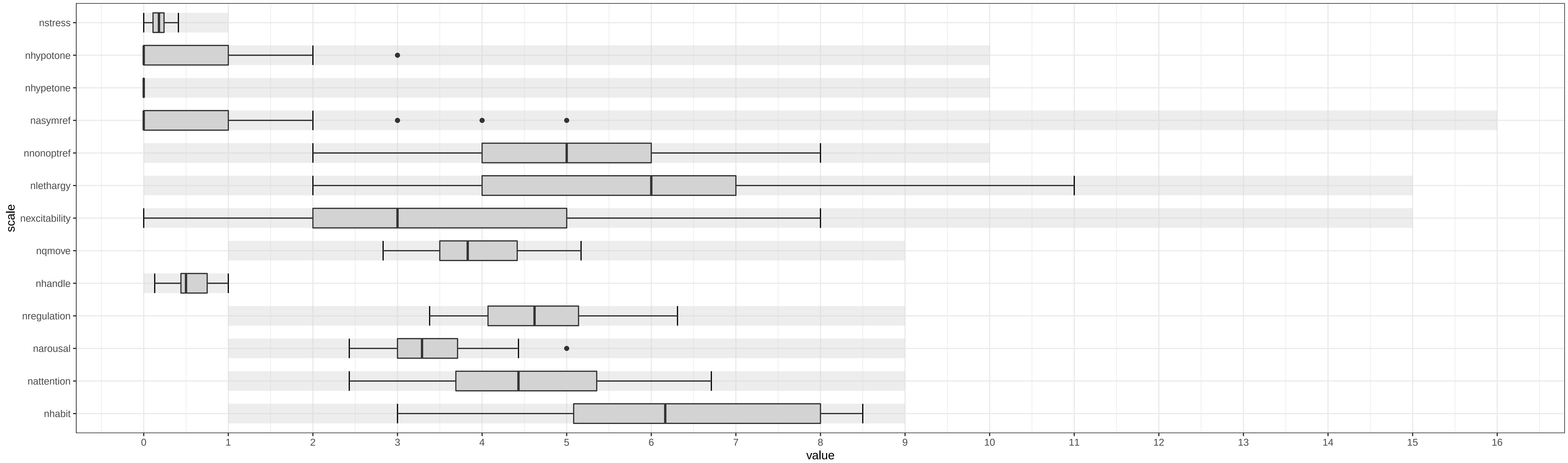
